# Early and late onset preeclampsia associated with different placental pathology and clinical risk characteristics

**DOI:** 10.1101/2022.12.05.22282973

**Authors:** Peilin Zhang, Naureen Shama

## Abstract

**Background:** Preeclampsia is classified as early onset and late onset types with different clinical manifestation and pathogenesis. Placental pathology of preeclampsia has been largely based on description of the early onset type, and the morphologic features of placenta from late onset preeclampsia were similar to those of non-preeclamptic placentas.

**Objective:** To identify clinically relevant maternal and neonatal risk characteristics and placental pathology for early onset and late onset preeclampsia.

**Study design:** We have collected all placental pathology information as well as maternal and neonatal birth information from March 2020 to December 2021 including preeclampsia and non-preeclampsia patients. We compared preterm and term preeclampsia in regards to maternal and neonatal complication and placental pathology by using logistic regression models to determine the important clinical risk factors associated with preeclampsia and placental pathology.

**Results:** A total 3724 placentas including 614 placentas from preeclamptic and 3110 non-preeclamptic patients were studied. Preterm preeclampsia (<37 weeks) was similar to early onset preeclampsia (<34 weeks) in maternal and neonatal complications as well as placental pathology, and these features were statistically different from those from term preeclampsia. More male fetuses were associated with early onset preeclampsia and female fetuses associated with late onset term preeclampsia when compared to non-preeclamptic patients. Maternal race/ethnicity and marital status were associated with preeclampsia but this association was no longer significant after logistic regression analysis. Preterm preeclampsia was statistically associated with gestational diabetes, placental infarcts, intrauterine fetal growth restriction and fetal vascular malperfusion, whereas term preeclampsia was associated with decidual vasculopathy and maternal obesity.

**Conclusions:** Early onset preeclampsia is a different clinical syndrome from the late onset type with clinical implication of pathogenesis and management.

## Introduction

Preeclampsia is a clinical syndrome with heterogeneous manifestations and severity [1, 2]. There are different classification schemes to characterize preeclampsia based on the etiology and pathogenesis, and early onset preeclampsia appears to be a different disease from the late onset preeclampsia in pathogenesis, clinical manifestation and severity as well as placental pathology [3, 4]. The placental pathology of preeclampsia has been reasonably consistent throughout most studies over time, and these morphologic features were mostly associated with early onset preeclampsia, although significant overlapping of placental pathology existed with late onset preeclampsia [5-7]. All morphologic features of the placental pathology of preeclampsia point to the disturbance of maternal placental circulation, and the features of abnormality of fetal placental circulation are not commonly identified [6-8]. In the latest guideline for placental pathology, the terminology adopted for clinical application was “maternal vascular malperfusion” which represents the constellation of a variety of morphologic descriptions in placental pathology literature including placental infarcts, intervillous thrombus, maternal decidual vasculopathy, villous maturation abnormalities (accelerated maturation) and placental abruption, the most lethal form of placental pathology in preeclampsia [6, 9, 10]. However, like many clinical features of preeclampsia, the morphologic features of placental pathology are not sufficiently specific to preeclampsia, and most of these morphologic characteristics can be seen in other maternal conditions in pregnancy, such as gestational diabetes, maternal obesity and advanced maternal ages [11, 12]. In clinical setting, many confounding factors invariably affect uteroplacental circulation, leading ultimately to various morphologic features of placental pathology. These confounding factors include maternal race and ethnicity, marital status, maternal obesity, advanced maternal age, gestational diabetes and other metabolic syndromes [13, 14]. In current study, we have chronologically collected maternal, neonatal and placental pathology information of pregnancy in our institution, and we used statistical and logistic regression models to determine the important factors for the associated risk of clinical preeclampsia and placental pathology.

## Materials and methods

The study was approved by the Institutional Review Board (IRB) at New York Presbyterian – Brooklyn Methodist Hospital, Brooklyn, NY ([1592673-1]). Placental examination in our institution is criteria-based, and the placentas submitted for pathology examination for a variety of clinical indications in March 2020 and December 2021 were included in the study. Routine paraffin-embedded tissues and Hematoxylin & Eosin (H&E) stained slides were examined by light microscopy using the Amsterdam criteria for placental examination [6]. The placental pathology data were entered into Excel spreadsheet (Microsoft Corporation) at the time of pathology examination, and the neonatal birth data including sex, birth weight, birth length and head circumference were subsequently retrieved from the medical records. Maternal racial data were retrieved from the medical record according to the US census criteria as Asians, non-Hispanic Black, Hispanic, and non-Hispanic white. Our racial data also included “others”, “unknown”, or “declined” as one group. Marital status was listed as married, single, divorced, life partner, or others including unknowns and declined to respond. Clinical complications and placental pathological findings were recorded as present or absent. Placental inflammatory / infectious conditions were not graded or staged, and all placental inflammatory responses were included. Only singleton placentas from third trimesters were included in the data, and placentas from first and second trimesters were excluded. Lab tests of white blood counts with differentials and blood pressures measurements were from pre-admission test before delivery only, and after delivery blood tests and blood pressure measurements were excluded. Statistical analysis was performed by using various programs of R-Package including baseline characteristic table, multivariate tests and logistic regression models (http://statistics4everyone.blogspot.com/2018/01/.html).

## Results

### 1 Early and late onset preeclampsia versus preterm and term preeclampsia

A total 3724 singleton placentas were included in current study including 614 (16.5%) placentas from patients with preeclampsia/pregnancy induced hypertension (PRE/PIH) and 3110 placentas from other clinical conditions that met the criteria for placental pathology examination. There were 74 placentas from early onset preeclampsia (defined as less than 34 weeks), representing 12.1 % of total preeclamptic patients, and 540 placentas from late onset preeclampsia (34 weeks or later), 87.9% of total preeclamptic patients. Maternal, neonatal and placental characteristics of early onset and late onset preeclampsia were significantly different on all categories examined as previously described (data not shown) [4, 15]. Late onset preeclampsia was further divided as “preterm late onset” (34 to 36 weeks) and “term late onset” preeclampsia, and maternal, neonatal and placental pathology features were compared to those of early onset preeclampsia (Table 1). There were 121 preterm preeclamptic placentas by combining the early onset preeclamptic placentas (N=74) and preterm late onset preeclamptic placentas (N=47) and 493 term preeclamptic placentas (Table 1). Statistically significant differences were observed in maternal, neonatal and placental pathology categories between the early onset preeclampsia and term late onset preeclampsia, and the characteristics of preterm late onset preeclampsia were in between the early onset and term late onset preeclampsia (Table 1). Statistically significantly more male fetuses were associated with early onset preeclampsia while more female fetuses were associated with late onset preeclampsia compared to non-preeclamptic patients (Table 1). Neonatal birth weight, birth length, head circumference and umbilical cord length were all statistically less for preterm preeclampsia than those of term preeclampsia (Table 1). Maternal race/ethnicity, marital status, and obesity were all associated with preeclampsia as previously described [13, 14]. There were significantly more Cesarean section delivery for preterm preeclampsia in comparison with late onset preeclampsia or non-preeclamptic patients (Table 1). Maternal vascular malperfusion (MVM), the characteristic and constellation of placental pathology features including the most lethal form of pathology changes, placental abruption, was predominantly found in early onset preeclampsia (Table 1). Maternal, neonatal and placental pathology characteristics of preterm preeclampsia and term preeclampsia were statistically significantly different in all categories examined, and these differences were similar, if not identical, to those between the early onset and late onset preeclampsia (data not shown).

**Table 1:**
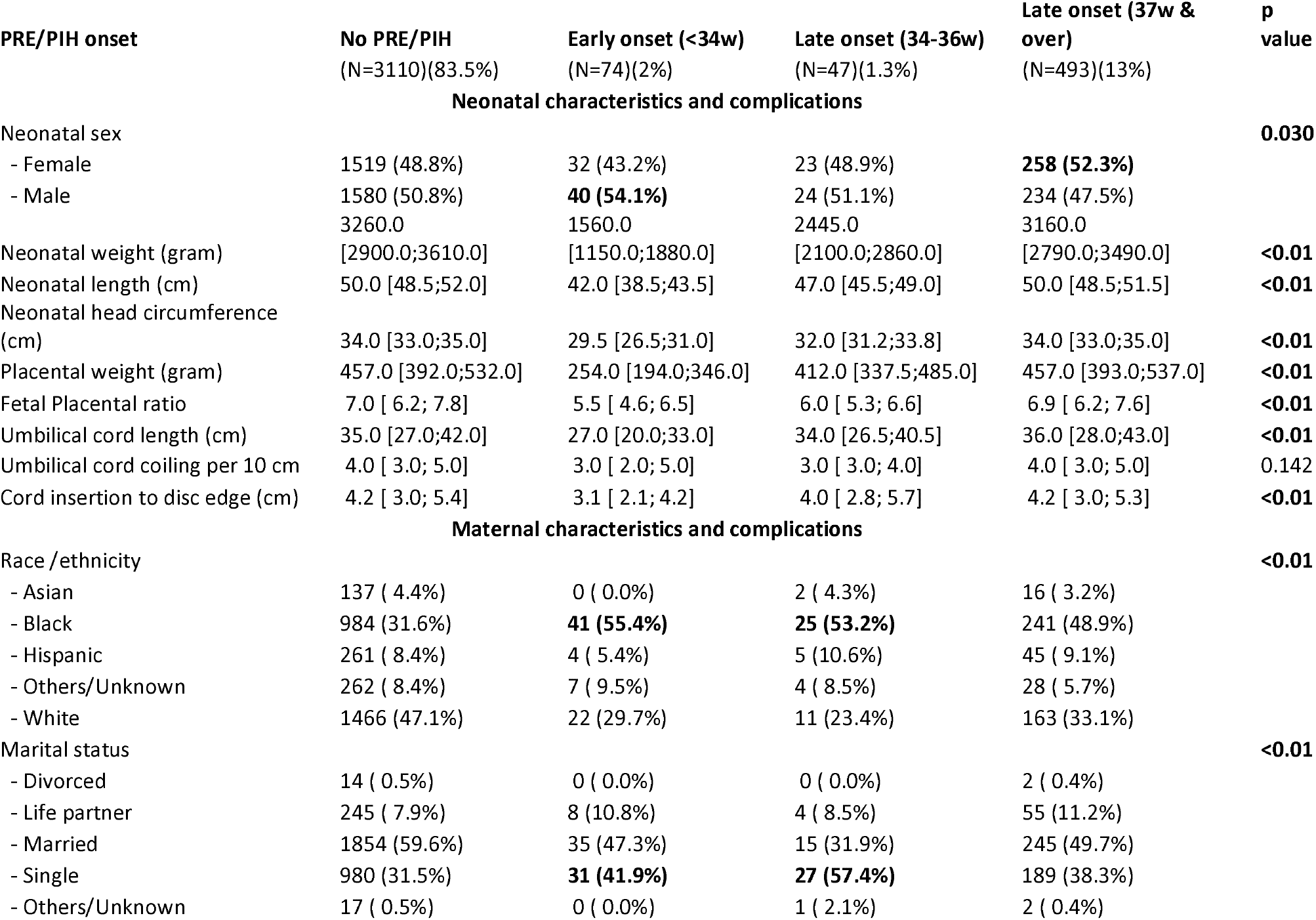
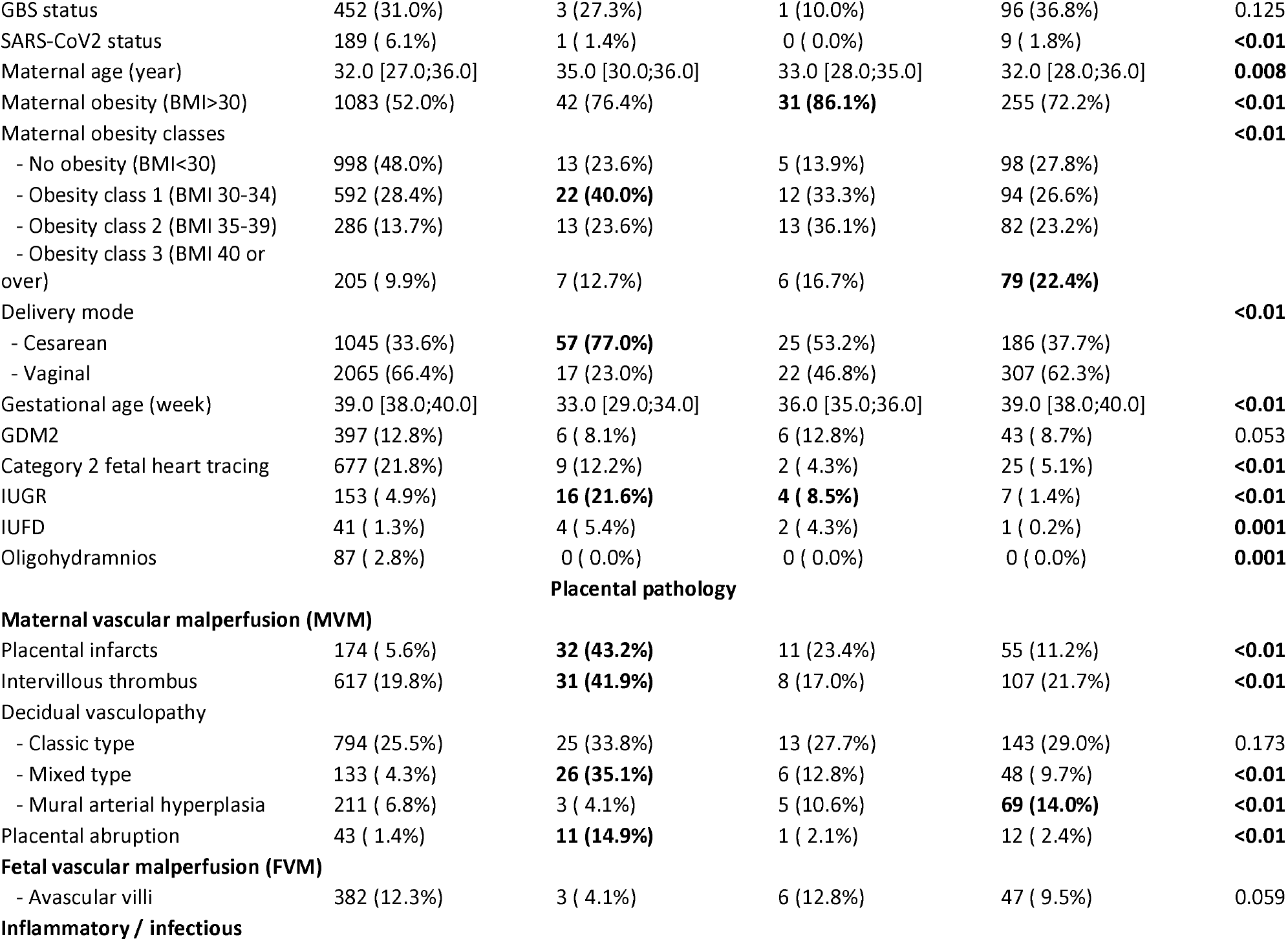
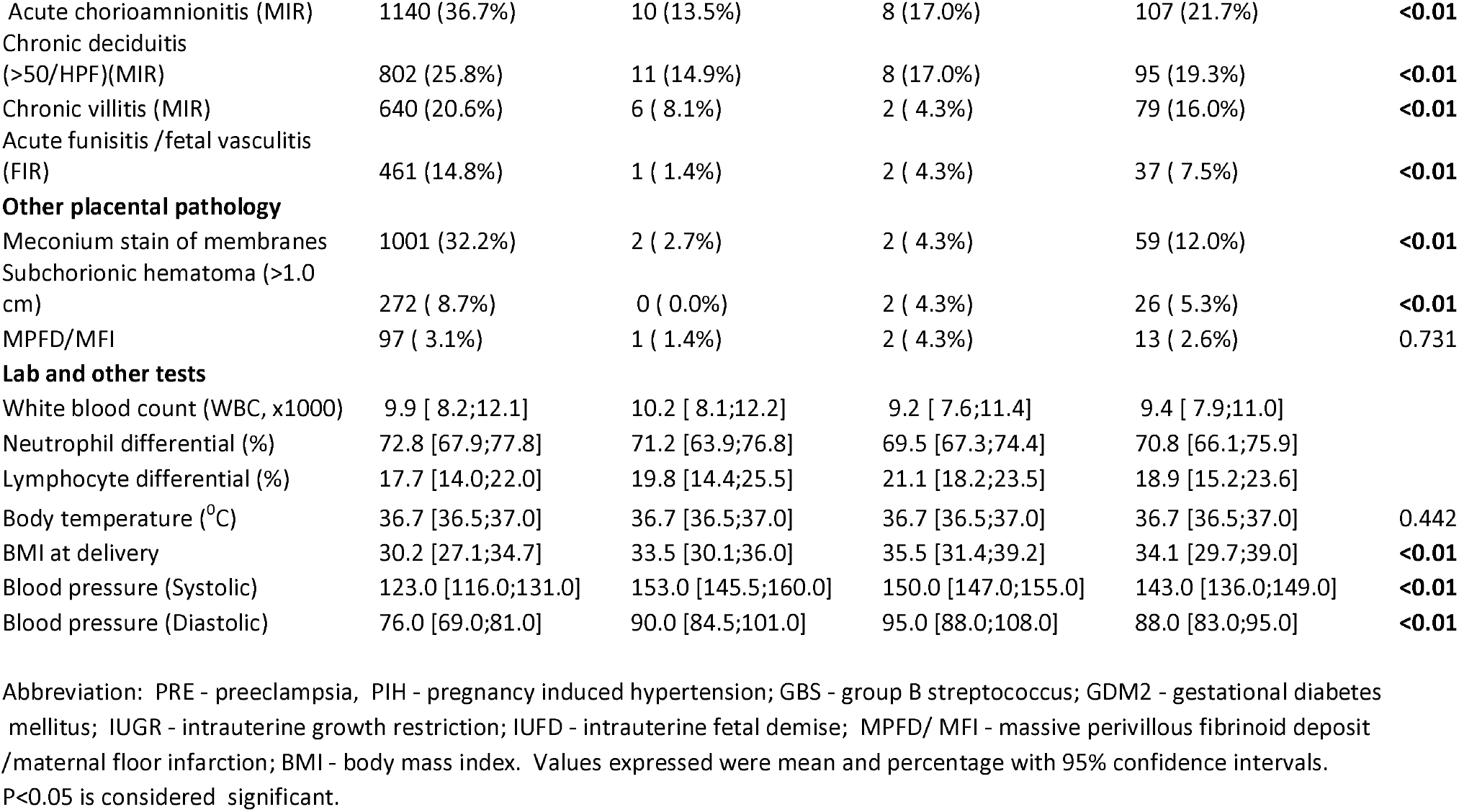
Comparison of preeclampsia onset with clinical pathologic features.

### 2 Logistic regression models and clinical features of preeclampsia with placental pathology

Maternal, neonatal and placental pathology features were examined by using Gaussian regression models for all preeclamptic patients including early onset and late onset preeclampsia and non-preeclamptic patients (Figure 1). Only two placental pathology features, namely mixed type vasculopathy and maternal obesity classes (class 1, class 2 and class 3) were statistically significantly associated with preeclampsia including early and late onset types (Figure 1). Other maternal, neonatal and placental pathology features were not associated with preeclampsia after excluding the confounding factors using logistic regression models. Similarly, GDM2, IUGR, placental infarcts and avascular villi (FVM) were found to be statistically significantly associated with preterm preeclampsia including early onset preeclampsia (Figure 2), whereas decidual vasculopathy including mixed type and classic type vasculopathy and maternal obesity classes were associated with term preeclampsia (Figure 3). The data from early onset preeclampsia versus late onset preeclampsia were similar to those described for preterm versus term preeclampsia (data not shown). Table 2 summarized the significant associations between preeclampsia types based on the onset and other maternal, neonatal and placental pathology features using Gaussian regression models.

**Table 2:** Significant clinical and pathological features associated with preeclampsia.

| **Total preeclampsia** | **p value** | **Preterm onset** | **p value** | **Term onset** | **p value** |
| --- | --- | --- | --- | --- | --- |
| Mixed type vasculopathy | <0.01 | GDM2 | 0.047 | Mixed type vasculopathy | <0.001 |
| Maternal obesity class | 5.30E-07 | IUGR | 0.019 | Maternal obesity classes | 7.68E-08 |
|  |  | Infarcts | 0.005 | Classic type vasculopathy | 0.029 |
|  |  | FVM (avascular villi) | 0.038 |  |  |

**Figure 1.**
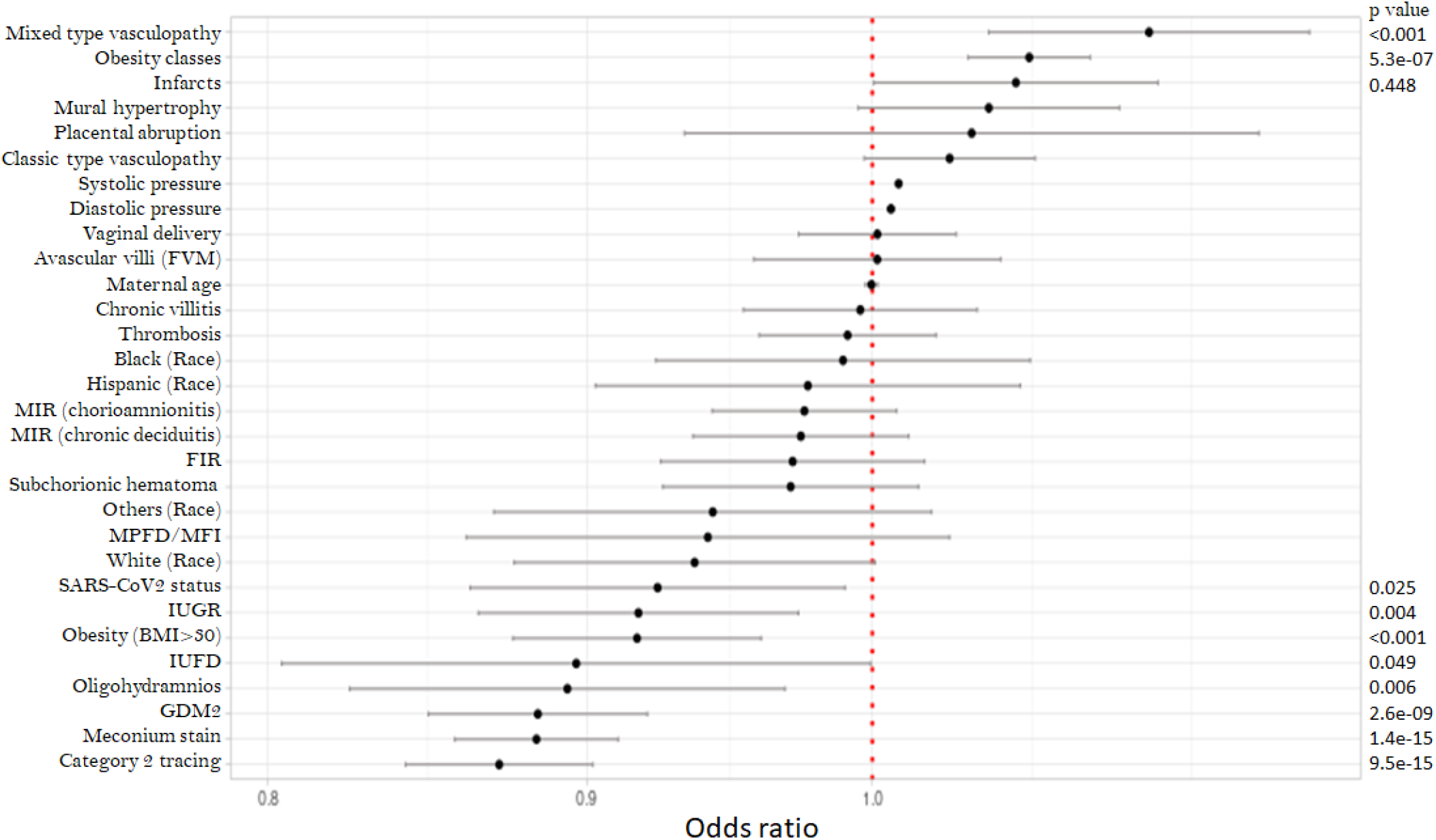
Logistic regression models of all preeclampsia (N=614) and non-preeclampsia placentas (N=3110) with associated maternal and neonatal complications. P<.05 is considered significant.

**Figure 2.**
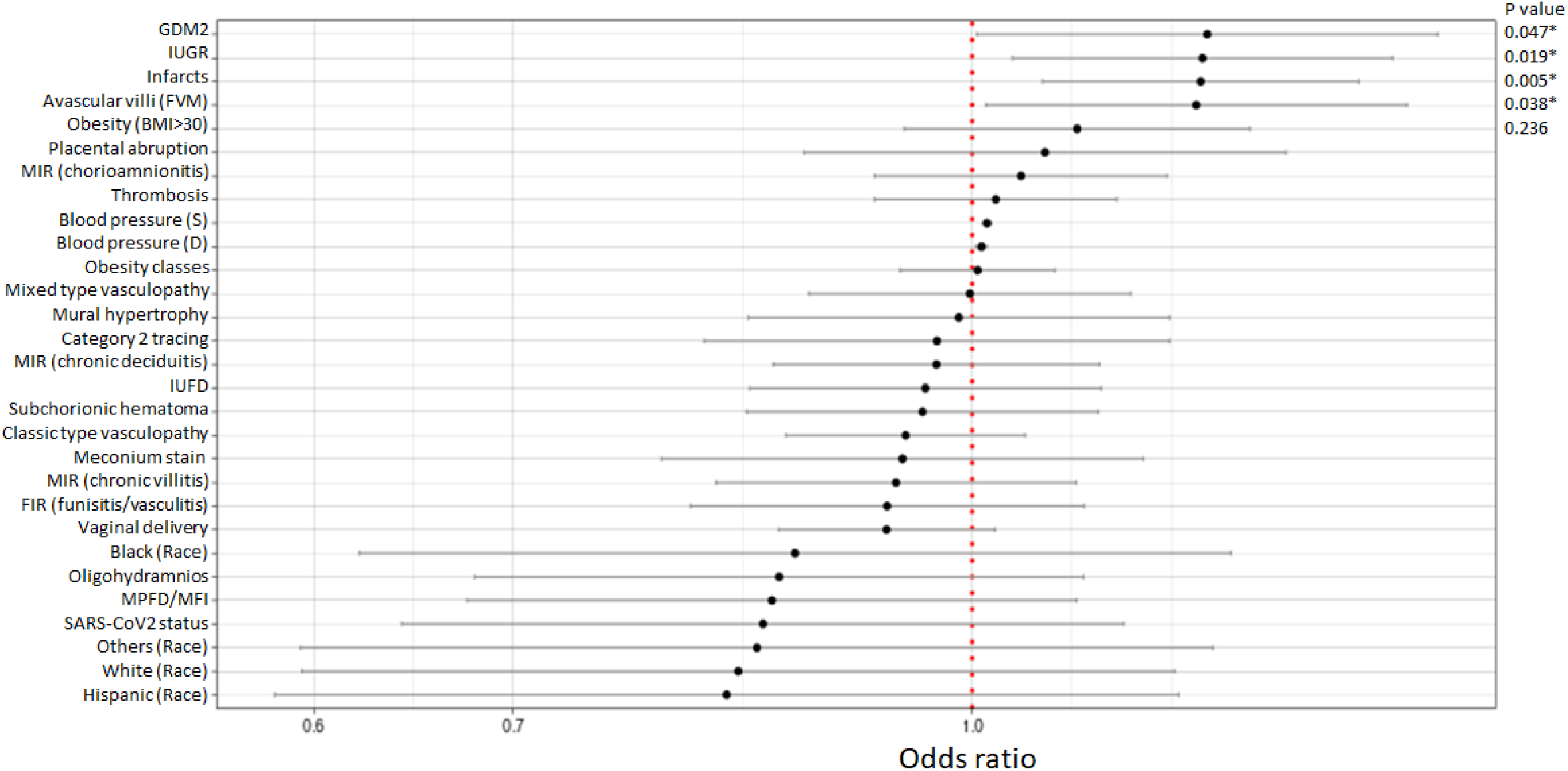
Logistic regression models of preterm preeclampsia (N=121) and non-preeclampsia placentas (N=295) with associated maternal and neonatal complications. P<.05 is considered significant

**Figure 3.**
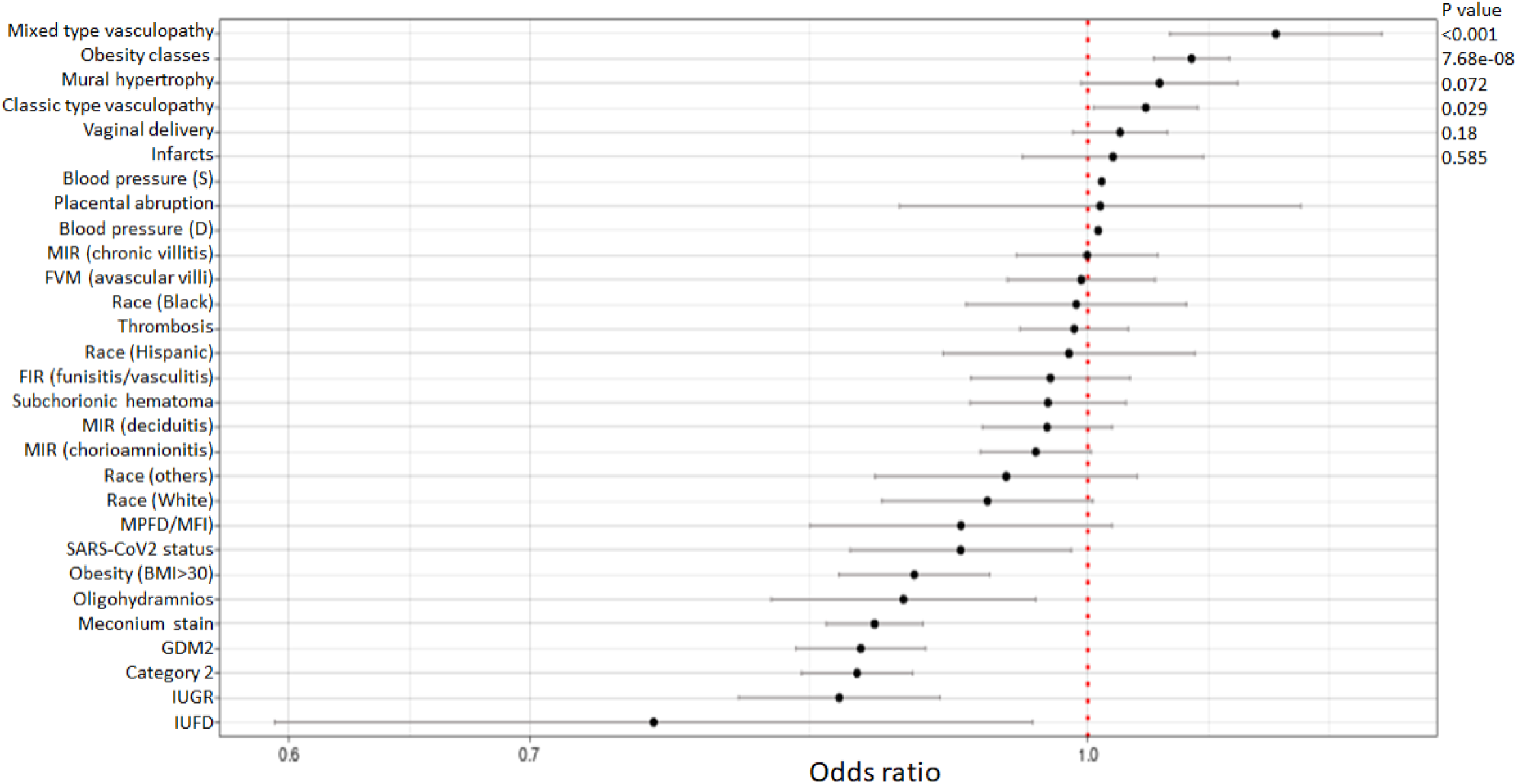
Logistic regression models of term preeclampsia (N=493) and non-preeclampsia placentas (N=2814) with associated maternal and neonatal complications. P<.05 is considered significant.

### 3 Preeclampsia and IUGR

There were totally 180 placentas with IUGR in our study population (180/3724, 4.8%), and 27 placentas were from the patients with preeclampsia (27/614, 4.4%) (Table 1). The incidence of IUGR in preeclampsia was significantly higher in early onset preeclampsia (16/74, 21.6%), and lower in late onset preeclampsia (11/540, 2%). Only 7 IUGR placentas were found in term preeclamptic patients (7/493, 1.4%), and this incidence was lower than that of non-preeclamptic patients (153/3110, 4.9%). Logistic regression model showed that IUGR was significantly associated with massive peri-villous fibrinoid deposit/maternal floor infarction (MPFD/MFI) and placental infarcts but not with preeclampsia overall (Table 4). IUGR was found to be associated with early onset type or preterm preeclampsia (Figure 2).

### 4 Preeclampsia and diabetes

A total 452 placentas were from gestational diabetic (GDM2) mothers including 25 placentas from preterm delivery (<37 weeks), and 427 from term delivery. Overall, gestational diabetes (GDM2) was statistically significant for increased neonatal weight, placental weight and the rate of macrosomia (>=4000 grams) (Supplementary Table 1) as previously known. Importantly, GDM2 overall was statistically associated with decreased, but not increased, frequency of preeclampsia (p=0.01) (Supplementary Table 1). When GDM2 was analyzed in preterm pregnancy (N=416), GDM2 was trending towards increased frequency of preeclampsia but statistically not significant (p=0.06) (Table 3), whereas GDM2 was negatively associated with preeclampsia in term pregnancy (N=3307)(Supplementary Table 2).

**Table 3:** GDM2 and Preterm Pregnancy (N=416)

| Preterm pregnancy (<37 w) | No GDM2 (N=391) | GDM2 (N=25) | Total (N=416) | p value |
| --- | --- | --- | --- | --- |
| **Neonatal birth data** |  |  |  |  |
| Neonatal sex |  |  |  | 0.25 |
| - Female | 178 (45.5%) | 8 (32.0%) | 186 (44.7%) |  |
| - Male | 203 (51.9%) | 17 (68.0%) | 220 (52.9%) |  |
| Neonatal weight (gram) | 2150.0 [1400.0;2550.0] | 2360.0 [2130.0;3010.0] | 2175.0 [1455.0;2575.0] | **<0.001** |
| Neonatal length (cm) | 45.0 [40.0;47.5] | 46.5 [44.5;49.0] | 45.0 [41.0;47.5] | **<0.001** |
| Neonatal head circumference (cm) | 31.0 [27.5;32.5] | 32.5 [31.0;34.0] | 31.0 [28.0;32.5] | **<0.001** |
| Placental weight (gram) | 331.0 [239.5;413.0] | 396.0 [354.0;456.0] | 336.0 [245.5;415.5] | **<0.001** |
| Fetal placental ratio (FPR) | 5.8 [ 4.8; 6.8] | 6.2 [ 5.8; 6.9] | 5.9 [ 4.9; 6.8] | **0.03** |
| Umbilical cord length (cm) | 29.0 [23.0;36.0] | 35.0 [26.0;41.0] | 29.0 [23.0;36.0] | 0.10 |
| Umbilical cord coiling per 10 cm | 3.0 [ 2.0; 5.0] | 3.0 [ 3.0; 6.0] | 3.0 [ 2.0; 5.0] | 0.38 |
| **Maternal characteristics and complications** |  |  |  |  |
| Race/ethnicity |  |  |  | **0.05** |
| - Asian | 4 ( 1.0%) | 2 ( 8.0%) | 6 ( 1.4%) |  |
| - Black | 183 (46.8%) | 11 (44.0%) | 194 (46.6%) |  |
| - Hispanic | 33 ( 8.4%) | 3 (12.0%) | 36 ( 8.7%) |  |
| - Others/unknown/declined | 48 (12.3%) | 4 (16.0%) | 52 (12.5%) |  |
| - White | 123 (31.5%) | 5 (20.0%) | 128 (30.8%) |  |
| Marital status |  |  |  | 0.85 |
| - Divorced | 1 ( 0.3%) | 0 ( 0.0%) | 1 ( 0.2%) |  |
| - Life partner | 34 ( 8.7%) | 3 (12.0%) | 37 ( 8.9%) |  |
| - Married | 189 (48.3%) | 14 (56.0%) | 203 (48.8%) |  |
| - Others/unknown/declined | 2 ( 0.5%) | 0 ( 0.0%) | 2 ( 0.5%) |  |
| - Single | 165 (42.2%) | 8 (32.0%) | 173 (41.6%) |  |
| Maternal age (year) | 32.0 [27.5;35.0] | 32.0 [29.0;38.0] | 32.0 [28.0;35.0] | 0.16 |
| Gestational age (week) | 34.0 [30.0;36.0] | 34.0 [33.0;36.0] | 34.0 [30.5;36.0] | 0.21 |
| Body mass index at delivery (BMI) | 30.5 [26.1;35.5] | 38.6 [32.5;40.8] | 30.8 [26.1;35.9] | **0.01** |
| Maternal obesity (BMI>30) | 157 (54.9%) | 13 (86.7%) | 170 (56.5%) | **0.03** |
| Obeisty classes |  |  |  | **0.03** |
| - No obesity | 129 (45.1%) | 2 (13.3%) | 131 (43.5%) |  |
| - Obesity class I (BMI 30-34) | 80 (28.0%) | 4 (26.7%) | 84 (27.9%) |  |
| - Obesity class II (BMI 35-39) | 38 (13.3%) | 4 (26.7%) | 42 (14.0%) |  |
| - Obesity class III (BMI 40 or over) | 39 (13.6%) | 5 (33.3%) | 44 (14.6%) |  |
| Delivery mode |  |  |  | **<0.001** |
| - Cesarean | 156 (39.9%) | 18 (72.0%) | 174 (41.8%) |  |
| - Vaginal | 235 (60.1%) | 7 (28.0%) | 242 (58.2%) |  |
| GBS status | 32 (36.4%) | 2 (33.3%) | 34 (36.2%) | 1.00 |
| **PRE/PIH** | **109 (27.9%)** | **12 (48.0%)** | **121 (29.1%)** | **0.06** |
| SARS-CoV2 status | 12 ( 3.1%) | 0 ( 0.0%) | 12 ( 2.9%) | 0.79 |
| IUGR | 29 ( 7.4%) | 0 ( 0.0%) | 29 ( 7.0%) | 0.31 |
| IUFD | 41 (10.5%) | 0 ( 0.0%) | 41 ( 9.9%) | 0.17 |
| Category 2 fetal tracing | 28 ( 7.2%) | 3 (12.0%) | 31 ( 7.5%) | 0.62 |
| Oligohydramnios | 12 ( 3.1%) | 1 ( 4.0%) | 13 ( 3.1%) | 1.00 |
| **Placental pathology** |  |  |  |  |
| **Maternal vascular malperfusion** |  |  |  |  |
| Decidual vasculopathy |  |  |  |  |
| - Classic type | 117 (29.9%) | 6 (24.0%) | 123 (29.6%) | 0.69 |
| - Mixed type | 53 (13.6%) | 6 (24.0%) | 59 (14.2%) | 0.25 |
| - Mural arterial hypertrophy | 18 ( 4.6%) | 6 (24.0%) | 24 ( 5.8%) | **<0.001** |
| Placental infarcts | 58 (14.8%) | 2 ( 8.0%) | 60 (14.4%) | 0.52 |
| Maternal thrombosis | 92 (23.5%) | 5 (20.0%) | 97 (23.3%) | 0.87 |
| Placental abruption | 20 ( 5.1%) | 0 ( 0.0%) | 20 ( 4.8%) | 0.50 |
| **Fetal vascular malperfusion (FVM)** | 35 ( 9.0%) | 2 ( 8.0%) | 37 ( 8.9%) | 1.00 |
| **Inflammtory/Infectious** |  |  |  |  |
| Acute chorioamnionitis (MIR) | 131 (33.5%) | 10 (40.0%) | 141 (33.9%) | 0.66 |
| Chronic deciduitis (>50/HPF)(MIR) | 74 (18.9%) | 6 (24.0%) | 80 (19.2%) | 0.72 |
| Chronic villitis (MIR) | 54 (13.8%) | 4 (16.0%) | 58 (13.9%) | 0.99 |
| Acute funisitis/fetal vasculitis (FIR) | 58 (14.8%) | 4 (16.0%) | 62 (14.9%) | 1.00 |
| **Other placental pathology** |  |  |  |  |
| Meconium stain of fetal membranes | 23 ( 5.9%) | 1 ( 4.0%) | 24 ( 5.8%) | 1.00 |
| Subchorionic hematoma (>1.0 cm) | 28 ( 7.2%) | 2 ( 8.0%) | 30 ( 7.2%) | 1.00 |
| MPFD/MFI | 9 ( 2.3%) | 1 ( 4.0%) | 10 ( 2.4%) | 1.00 |
| **Lab and other tests** |  |  |  |  |
| WBC count x1000 | 10.6 [ 8.2;12.8] | 10.0 [ 6.8;12.0] | 10.6 [ 8.1;12.8] | 0.21 |
| Neutrophil differential (%) | 72.2 ± 8.0 | 72.6 ± 6.7 | 72.3 ± 8.0 | 0.86 |
| Lymphocyte differential (%) | 18.6 [14.2;22.8] | 18.6 [14.9;22.8] | 18.6 [14.3;22.8] | 0.92 |
| Body temperature (°C) | 36.8 [36.5;37.0] | 36.7 [36.6;37.0] | 36.8 [36.5;37.0] | 0.85 |
| Blood pressure (Systolic) | 128.0 [118.5;143.5] | 146.0 [118.5;152.0] | 129.5 [118.5;146.0] | 0.16 |
| Blood pressure (Diastolic) | 79.5 [69.5;88.0] | 86.5 [69.0;96.5] | 80.0 [69.5;88.0] | 0.24 |

Abbreviation: PRE - preeclampsia, PIH - pregnancy induced hypertension; GBS - group B streptococcus; GDM2 - gestational diabetes mellitus; IUGR - intrauterine growth restriction; IUFD - intrauterine fetal demise; MPFD/ MFI - massive perivillous fibrinoid deposit /maternal floor infarction; BMI - body mass index. Values expressed were mean and percentage with 95% confidence intervals. P<0.05 is considered significant.

Gaussian regression model analysis of GDM2 revealed that chronic villitis was significantly associated with GDM2 positively overall (Supplementary Figure 1), and this positive association between GDM2 and chronic villitis was found in term pregnancy but not in preterm pregnancy (Supplementary Figure 2), whereas GDM2 was significantly associated with mural arterial hypertrophy and preeclampsia in preterm pregnancy (Table 4).

**Table 4:** Significant clinical and pathologic features associated with IUGR and GDM2.

| **IUGR** | **p**<br>**value** | **Total GDM2** | **p**<br>**value** | **Preterm GDM2** | **p**<br>**value** | **Term GDM2** | **p**<br>**value** |
| --- | --- | --- | --- | --- | --- | --- | --- |
| MPFD/MFI | 0.046 | MIR (chronic villitis) | 0.027 | Mural hypertrophy | 0.031 | MIR (chronic villitis) | 0.015 |
| Placental infarcts | 0.004 | Obesity | 0.154 | **PRE/PIH** | 0.047 |  |  |
|  |  |  |  | MIR (chorioamnionitis) | 0.051 |  |  |

There were 55 patients with combined GDM2 and preeclampsia (Supplementary Table 3). Combination of GDM2 and preeclampsia showed statistically significant association with increased Cesarean section rate, maternal obesity class III, mixed type vasculopathy, placental infarcts and placental abruption compared to GDM2 or preeclampsia alone.

## Discussion

Our current data showed that statistically significant differences were observed between the early onset preeclampsia and late onset type based on the gestational age of 34 weeks, and these differences were identified in all categories of maternal and neonatal complications and placental pathology. The difference between early onset and late onset types based on gestational age of 34 weeks was similar to those between preterm and term preeclampsia based on 37 weeks. Our data also showed that term preeclampsia was significantly associated with decidual vasculopathy and maternal obesity, whereas preterm preeclampsia was associated with GDM2, IUGR, placental infarcts and fetal vascular malperfusion (FVM, avascular villi). Decidual vasculopathy was classified as three different types, classic type, mixed type and mural arterial hypertrophy/hyperplasia, and these three types of decidual vasculopathy and their clinical significance were previously described [23]. Classic type vasculopathy including atherosis of macrophage type and atherosis of trophoblastic type was shown to be associated with preeclampsia [23, 24]. Atherosis of trophoblast type depicted the failure of trophoblast cell death (fail to die) or the failure of endothelial regeneration (fail to regenerate) of remodeled spiral artery, which occurs in late pregnancy (third trimester) [25, 26]. It is interesting to note that early onset or preterm preeclampsia was not associated with decidual vasculopathy in our data, and our previous study showed that mixed type vasculopathy was associated with lowest neonatal birth weight and placental weight [21, 23]. Maternal obesity and advanced maternal age were associated with significant clinical complications and placental pathology [13, 14]. However, a number of clinical and pathologic factors such as non-Hispanic black race/ethnicity, intervillous thrombus, placental inflammatory response (both MIR and FIR) previously demonstrated as risk factors to preeclampsia were no longer significant after logistic regression model analysis. Interestingly, fetal vascular malperfusion (avascular villi) was found associated with preterm preeclampsia but not with term preeclampsia. FVM was known to be associated with fetal thromboembolic conditions including Factor V Leiden deficiency, cord obstruction, fetal vascular thrombosis, IUGR or stillbirth [8, 27-30]. Only one study previously demonstrated the association of FVM with preterm preeclampsia [8]. Our results and other’s demonstrate that preterm preeclampsia characterized by maternal vascular malperfusion can also affect fetal placental circulation, leading to fetal vascular malperfusion through unknown mechanisms [8]. Finally, our data confirmed that IUGR was associated with preterm preeclampsia and placental infarcts, but not with term preeclampsia or other placental pathology except for massive peri-villous fibrinoid deposit /maternal floor infarction (MPFD/MFI). IUGR was known to be associated with MPFD/MFI for unclear mechanisms [31, 32].

Preeclampsia has been classified as early onset and late onset types based on the gestational ages of clinical manifestations, and preeclampsia has been extensively studied overall in maternal, and neonatal complications as well as placental pathology [1]. It is generally believed that early onset preeclampsia is a different disease with deficiency of spiral artery remodeling, leading to placental malperfusion (underperfusion) whereas the late onset preeclampsia was due to maternal systemic conditions [3]. Both early onset and late onset preeclampsia showed placental malperfusion (underperfusion) characterized by abnormal serum angiogenic factor levels, although the size and weight of placentas from late onset preeclampsia were not strikingly different from the non-preeclamptic patients [16-22]. Traditional descriptions of clinical manifestations and placental pathology of preeclampsia or eclampsia were largely based on the characteristics of early onset type, although significant maternal and neonatal complications as well as placental pathology can occur in late onset preeclampsia [1].

The relationship between diabetes and preeclampsia is more complex [33, 34]. Our data showed that late onset and term preeclampsia was negatively associated with GDM2, and early onset preeclampsia was not statistically associated with GDM2. However, logistic regression analysis revealed statistically positive association between preterm preeclampsia with GDM2. Moreover, combination of GDM2 and preeclampsia increased risks of Cesarean section, mixed type decidual vasculopathy, placental infarcts and placental abruption in comparison to preeclampsia or GDM2 alone, and these patients were associated with more advanced maternal obesity (class III).

Our data demonstrated that early onset and late onset preeclampsia were different in clinical risk features and placental pathology. Separating the two different types of preeclampsia in early second trimester will benefit clinical monitoring of the patients and the risk assessment of maternal and neonatal complications. Late onset or term preeclampsia will be difficult to differentiate from normal pregnancy in second trimester as clinical and placental pathology features were similar if not identical. Our current data revealed not only different placental pathology features between the early onset preeclampsia and late onset type, but also the clinical risk characteristics using statistical models. It seems that the two stage theory of preeclampsia fit more into early onset preeclampsia which represents small percentage of patients with severe maternal and neonatal syndromes [35-37]. Late onset preeclampsia appears more metabolically driven and it is associated with maternal obesity and decidual vasculopathy not related to deficient spiral artery remodeling in early pregnancy.

Our study demonstrated clearly that early onset preeclampsia was different from late onset type in clinical risk characteristics and placental pathology features excluding many confounding factors using statistical and logistic regression models. Our study also showed sex dimorphism of early onset and late onset preeclampsia with unclear molecular mechanisms. Our study also demonstrated that early onset preeclampsia was associated with fetal vascular malperfusion (avascular villi) which has not been extensively studied previously. The data was from a single urban community hospital with similar demographics and access to healthcare. The limitation was that although the total number of patients in the study was adequate, the case number of preeclampsia was mainly late onset type (term preeclampsia, 87.9%) and early onset preeclampsia was still small (74 cases, 12.1%). The conclusions from the study should be interpreted with caution due to the sample size. The other limitation is that preeclampsia was not stratified with severity (preeclampsia with severe features), and it is unclear if preeclampsia with severe features was associated with more placental pathology (more quantitative) and more clinical risks.

## Conclusions

Early onset or preterm preeclampsia was shown to be a syndrome with significant risk factors different from those of late onset or term preeclampsia. Proper diagnosis and management of early onset preeclampsia will improve maternal and neonatal outcomes of preeclampsia.

## Data Availability

All data produced in the present work are contained in the manuscript.

## Acknowledgements

None.

## Figure legend

Supplementary Figure 1: Logistic regression models of GDM2 (N=452) and non-GDM2 placentas (N=3272) with associated maternal and neonatal complications. P<.05 is considered significant.

Supplementary Figure 2. Logistic regression models of GDM2 (N=427) and non-GDM2 placentas in term pregnancy (N=2880) with associated maternal and neonatal complications. P<.05 is considered significant.

## Reference

1. Taylor, R., et al., Chesley’s Hypertensive Disorders in Pregnancy. 2015, London: Academic Press.

2. Irgens, H.U., et al., Long term mortality of mothers and fathers after pre-eclampsia: population based cohort study. BMJ, 2001. 323(7323): p. 1213–7.

3. Staff, A.C., R. Dechend, and C.W. Redman, Review: Preeclampsia, acute atherosis of the spiral arteries and future cardiovascular disease: two new hypotheses. Placenta, 2013. 34 Suppl: p. S73–8.

4. Burton, G.J., et al., Pre-eclampsia: pathophysiology and clinical implications. BMJ, 2019. 366: p. l2381.

5. Benirschke, K., G.J. Burton, and R.N. Baergen Pathology of the Human Placenta. 6th ed. 2012: Springer.

6. Khong, T.Y., et al., Sampling and Definitions of Placental Lesions: Amsterdam Placental Workshop Group Consensus Statement. Arch Pathol Lab Med, 2016. 140(7): p. 698–713.

7. Ernst, L.M., Maternal vascular malperfusion of the placental bed. APMIS, 2018. 126(7): p. 551–560.

8. Salafia, C.M., et al., Placental pathologic features of preterm preeclampsia. Am J Obstet Gynecol, 1995. 173(4): p. 1097–105.

9. Redline, R.W., et al., Maternal vascular underperfusion: nosology and reproducibility of placental reaction patterns. Pediatr Dev Pathol, 2004. 7(3): p. 237–49.

10. Redline, R.W., Placental pathology: Pathways leading to or associated with perinatal brain injury in experimental neurology, special issue: Placental mediated mechanisms of perinatal brain injury. Exp Neurol, 2022. 347: p. 113917.

11. Kim, Y.M., et al., Placental lesions associated with acute atherosis. J Matern Fetal Neonatal Med, 2015. 28(13): p. 1554–62.

12. Romero, R., et al., The frequency and type of placental histologic lesions in term pregnancies with normal outcome. J Perinat Med, 2018. 46(6): p. 613–630.

13. Zhang, P., et al., Placental pathology associated with maternal age and maternal obesity in singleton pregnancy. J Matern Fetal Neonatal Med, 2022: p. 1–10.

14. Zhang, P., et al., Differences in Prevalence of Pregnancy Complications and Placental Pathology by Race and Ethnicity in a New York Community Hospital. JAMA Netw Open, 2022. 5(5): p. e2210719.

15. Stepan, H., M. Hund, and T. Andraczek, Combining Biomarkers to Predict Pregnancy Complications and Redefine Preeclampsia: The Angiogenic-Placental Syndrome. Hypertension, 2020. 75(4): p. 918–926.

16. Soto, E., et al., Late-onset preeclampsia is associated with an imbalance of angiogenic and antiangiogenic factors in patients with and without placental lesions consistent with maternal underperfusion. J Matern Fetal Neonatal Med, 2012. 25(5): p. 498–507.

17. Triunfo, S., et al., Association of first-trimester angiogenic factors with placental histological findings in late-onset preeclampsia. Placenta, 2016. 42: p. 44–50.

18. Crovetto, F., et al., First-trimester screening with specific algorithms for early- and late-onset fetal growth restriction. Ultrasound Obstet Gynecol, 2016. 48(3): p. 340–8.

19. Baltajian, K., et al., Placental lesions of vascular insufficiency are associated with anti-angiogenic state in women with preeclampsia. Hypertens Pregnancy, 2014. 33(4): p. 427–39.

20. Rana, S., et al., Clinical characterization and outcomes of preeclampsia with normal angiogenic profile. Hypertens Pregnancy, 2013. 32(2): p. 189–201.

21. Zhang, P., Phenotypic Switch of Endovascular Trophoblasts in Decidual Vasculopathy with Implication for Preeclampsia and Other Pregnancy Complications. Fetal Pediatr Pathol, 2020: p. 1–20.

22. Karumanchi, S.A., Angiogenic Factors in Preeclampsia: From Diagnosis to Therapy. Hypertension, 2016. 67(6): p. 1072–9.

23. Zhang, P. and R. Baergen, Atherosis of Trophoblast Type. Arch Pathol Lab Med, 2022.

24. Zeek, P.M. and N.S. Assali, Vascular changes in the decidua associated with eclamptogenic toxemia of pregnancy. Am J Clin Pathol, 1950. 20(12): p. 1099–1109.

25. Zhang, P., Decidual Vasculopathy in Preeclampsia and Spiral Artery Remodeling Revisited: Shallow Invasion versus Failure of Involution. AJP Rep, 2018. 8(4): p. e241–e246.

26. Zhang, P., Decidual vasculopathy and spiral artery remodeling revisited II: relations to trophoblastic dependent and independent vascular transformation. J Matern Fetal Neonatal Med, 2020: p. 1–7.

27. Rogers, B.B., et al., Avascular villi, increased syncytial knots, and hypervascular villi are associated with pregnancies complicated by factor V Leiden mutation. Pediatr Dev Pathol, 2010. 13(5): p. 341–7.

28. Gibbins, K.J., et al., Stillbirth, hypertensive disorders of pregnancy, and placental pathology. Placenta, 2016. 43: p. 61–8.

29. Redline, R.W., et al., Fetal vascular obstructive lesions: nosology and reproducibility of placental reaction patterns. Pediatr Dev Pathol, 2004. 7(5): p. 443–52.

30. Heider, A., Fetal Vascular Malperfusion. Arch Pathol Lab Med, 2017. 141(11): p. 1484-1489.

31. Katzman, P.J. and D.R. Genest, Maternal floor infarction and massive perivillous fibrin deposition: histological definitions, association with intrauterine fetal growth restriction, and risk of recurrence. Pediatr Dev Pathol, 2002. 5(2): p. 159–64.

32. Andres, R.L., et al., The association of maternal floor infarction of the placenta with adverse perinatal outcome. Am J Obstet Gynecol, 1990. 163(3): p. 935–8.

33. Salzer, L., K. Tenenbaum-Gavish, and M. Hod, Metabolic disorder of pregnancy (understanding pathophysiology of diabetes and preeclampsia). Best Pract Res Clin Obstet Gynaecol, 2015. 29(3): p. 328–38.

34. Weissgerber, T.L. and L.M. Mudd, Preeclampsia and diabetes. Curr Diab Rep, 2015. 15(3): p. 9. 35.

35. Roberts, J.M. and C. Escudero, The placenta in preeclampsia. Pregnancy Hypertens, 2012. 2(2): p. 72–83.

36. Staff, A.C., The two-stage placental model of preeclampsia: An update. J Reprod Immunol, 2019. 134-135: p. 1–10.

37. Roberts, J.M. and C.A. Hubel, The two stage model of preeclampsia: variations on the theme. Placenta, 2009. 30 Suppl A: p. S32–7.

