## Supplemental tables and figures for "Early and late onset preeclampsia associated with different placental pathology and clinical risk characteristics": Supplementary material (Zhang Tables and Figures).docx

| **Supplementary table 1: GDM2 with clinical and neonatal features and placental pathology** | | | | |
| --- | --- | --- | --- | --- |
|  | **No GDM2 (N=3272)** | **GDM2 (N=452)** | **Total (N=3724)** | **p value** |
| **Neonatal birth data** | | | | |
| Neonatal sex |  |  |  | 0.37 |
| - Female | 1611 (49.2%) | 221 (48.9%) | 1832 (49.2%) |  |
| - Male | 1647 (50.3%) | 231 (51.1%) | 1878 (50.4%) |  |
| Neonatal weight (gram) | 3220.0 [2820.0;3560.0] | 3340.0 [2990.0;3670.0] | 3230.0 [2840.0;3580.0] | **<0.001** |
| Macrosomia (>=4000 g) | 225 ( 6.9%) | 53 (11.7%) | 278 ( 7.5%) | **<0.001** |
| Neonatal length (cm) | 50.0 [48.0;52.0] | 50.0 [49.0;52.0] | 50.0 [48.0;52.0] | **0.02** |
| Neonatal head circumference (cm) | 34.0 [33.0;35.0] | 34.0 [33.0;35.0] | 34.0 [33.0;35.0] |  |
| Placental weight (gram) | 451.0 [386.0;526.0] | 478.5 [413.5;557.5] | 454.0 [389.0;530.0] | **<0.001** |
| Fetal placental ratio (FPR) | 7.0 [ 6.2; 7.8] | 6.9 [ 6.1; 7.7] | 7.0 [ 6.2; 7.8] | 0.46 |
| Umbilical cord length (cm) | 34.0 [27.0;42.0] | 35.0 [27.0;43.0] | 35.0 [27.0;42.0] | 0.35 |
| Umbilical cord coiling per 10 cm | 4.0 [ 3.0; 5.0] | 4.0 [ 3.0; 5.0] | 4.0 [ 3.0; 5.0] | 0.85 |
| **Maternal characteristics and complications** | | | | |
| Race/ethnicity |  |  |  | **<0.001** |
| - Asian | 113 ( 3.5%) | 42 ( 9.3%) | 155 ( 4.2%) |  |
| - Black | 1163 (35.5%) | 128 (28.3%) | 1291 (34.7%) |  |
| - Hispanic | 262 ( 8.0%) | 53 (11.7%) | 315 ( 8.5%) |  |
| - Others/unknown/declined | 260 ( 7.9%) | 41 ( 9.1%) | 301 ( 8.1%) |  |
| - White | 1474 (45.0%) | 188 (41.6%) | 1662 (44.6%) |  |
| Marital status |  |  |  | 0.11 |
| - Divorced | 11 ( 0.3%) | 5 ( 1.1%) | 16 ( 0.4%) |  |
| - Life partner | 276 ( 8.4%) | 36 ( 8.0%) | 312 ( 8.4%) |  |
| - Married | 1874 (57.3%) | 275 (60.8%) | 2149 (57.7%) |  |
| - Others/unknown/declined | 8 ( 0.2%) | 0 ( 0.0%) | 8 ( 0.2%) |  |
| - Single | 1091 (33.3%) | 136 (30.1%) | 1227 (32.9%) |  |
| Delivery mode |  |  |  | 0.09 |
| - Cesarean | 1137 (34.7%) | 176 (38.9%) | 1313 (35.3%) |  |
| - Vaginal | 2135 (65.3%) | 276 (61.1%) | 2411 (64.7%) |  |
| Maternal age (year) | 32.0 [27.0;35.0] | 34.0 [30.0;37.0] | 32.0 [27.0;36.0] | **<0.001** |
| Gestational age (week) | 39.0 [38.0;40.0] | 39.0 [38.0;40.0] | 39.0 [38.0;40.0] |  |
| Preterm delivery (<37 week) | 391 (12.0%) | 25 ( 5.5%) | 416 (11.2%) | **<0.001** |
| GBS status | 476 (31.6%) | 76 (32.2%) | 552 (31.7%) | 0.92 |
| SARS-CoV2 status | 189 ( 5.8%) | 10 ( 2.2%) | 199 ( 5.3%) | **<0.001** |
| Body mass index at delivery (BMI) | 30.5 [27.3;35.2] | 32.7 [28.8;37.9] | 30.8 [27.4;35.6] | **<0.001** |
| Maternal obesity (BMI>30) | 1188 (54.2%) | 223 (67.0%) | 1411 (55.9%) | **<0.001** |
| Obesity classes |  |  |  | **<0.001** |
| - No obesity | 1004 (45.8%) | 110 (33.0%) | 1114 (44.1%) |  |
| - Obesity class I (BMI 30-34) | 612 (27.9%) | 108 (32.4%) | 720 (28.5%) |  |
| - Obesity class II (BMI 35-39) | 332 (15.1%) | 62 (18.6%) | 394 (15.6%) |  |
| - Obesity class III (BMI 40 or over) | 244 (11.1%) | 53 (15.9%) | 297 (11.8%) |  |
| **PRE/PIH** | **559 (17.1%)** | **55 (12.2%)** | **614 (16.5%)** | **0.01** |
| IUGR | 165 ( 5.0%) | 15 ( 3.3%) | 180 ( 4.8%) | 0.14 |
| IUFD | 48 ( 1.5%) | 0 ( 0.0%) | 48 ( 1.3%) | **0.02** |
| Category 2 fetal tracing | 685 (20.9%) | 28 ( 6.2%) | 713 (19.1%) | **<0.001** |
| Oligohydramnios | 83 ( 2.5%) | 4 ( 0.9%) | 87 ( 2.3%) | 0.04 |
| **Placental pathology** | | | | |
| **Maternal vascular malperfusion** |  |  |  |  |
| Decidual vasculopathy |  |  |  |  |
| - Classic type | 870 (26.6%) | 105 (23.2%) | 975 (26.2%) | 0.14 |
| - Mixed type | 186 ( 5.7%) | 27 ( 6.0%) | 213 ( 5.7%) | 0.89 |
| - Mural arterial hypertrophy | 234 ( 7.2%) | 54 (11.9%) | 288 ( 7.7%) | **<0.001** |
| Placental infarcts | 247 ( 7.5%) | 25 ( 5.5%) | 272 ( 7.3%) | 0.15 |
| Maternal thrombosis | 670 (20.5%) | 93 (20.6%) | 763 (20.5%) | 1.00 |
| Placental abruption | 63 ( 1.9%) | 4 ( 0.9%) | 67 ( 1.8%) | 0.17 |
| **Fetal vascular malperfusion (FVM)** | |  |  |  |
| - Avascular villi | 384 (11.7%) | 54 (11.9%) | 438 (11.8%) | 0.96 |
| **Inflammatory/Infectious** |  |  |  |  |
| MIR (acute chorioamnionitis) | 1152 (35.2%) | 113 (25.0%) | 1265 (34.0%) | **<0.001** |
| MIR (chronic deciduitis) | 797 (24.4%) | 119 (26.3%) | 916 (24.6%) | 0.39 |
| MIR (chronic villitis) | 619 (18.9%) | 108 (23.9%) | 727 (19.5%) | **0.02** |
| FIR (acute funisitis /fetal vasculitis) | 456 (13.9%) | 45 (10.0%) | 501 (13.5%) | **0.02** |
| **Other placental pathology** |  |  |  |  |
| Meconium stain of fetal membranes | 997 (30.5%) | 67 (14.8%) | 1064 (28.6%) | **<0.001** |
| Subchorionic hematoma (>1.0 cm) | 267 ( 8.2%) | 33 ( 7.3%) | 300 ( 8.1%) | 0.59 |
| MPFD/MFI | 106 ( 3.2%) | 7 ( 1.5%) | 113 ( 3.0%) | 0.07 |
| **Lab and other tests** |  |  |  |  |
| WBC count (x1000) | 9.9 [ 8.2;12.0] | 9.4 [ 7.7;11.1] | 9.8 [ 8.2;11.9] | **<0.001** |
| Neutrophil differential (%) | 72.7 [67.7;77.6] | 71.5 [67.4;75.9] | 72.5 [67.6;77.4] | **<0.001** |
| Lymphocyte differential (%) | 17.8 [13.9;22.1] | 19.3 [15.3;23.2] | 18.0 [14.2;22.3] | **<0.001** |
| Body temperature at admission (^0^C) | 36.7 [36.5;37.0] | 36.7 [36.5;37.0] | 36.7 [36.5;37.0] | 0.87 |
| Blood pressure (Systolic) | 126.0 [117.0;136.0] | 128.0 [120.0;135.0] | 126.0 [118.0;136.0] | 0.32 |
| Blood pressure (Diastolic) | 77.0 [70.0;85.0] | 79.0 [72.0;84.0] | 78.0 [70.0;84.0] | 0.12 |
| Abbreviation: PRE - preeclampsia, PIH - pregnancy induced hypertension; GBS - group B streptococcus; GDM2 - | | | | |
| gestational diabetes mellitus; IUGR - intrauterine growth restriction; IUFD - intrauterine fetal demise; | | | | |
| MPFD/ MFI - massive perivillous fibrinoid deposit /maternal floor infarction; BMI - body mass index. | | | |  |
| Values expressed were mean and percentage with 95% confidence intervals. P<0.05 is considered significant. | | | | |

| **Supplementary table 2: GDM2 in term pregnancy (N=3307)** | | | | |
| --- | --- | --- | --- | --- |
|  | **No GDM2 (N=2880)** | **GDM2 (N=427)** | **Total (N=3307)** | **p value** |
| **Neonatal birth data** | | | | |
| Neonatal sex |  |  |  | 0.55 |
| - Female | 1431 (49.7%) | 215 (50.4%) | 1646 (49.8%) |  |
| - Male | 1447 (50.2%) | 211 (49.4%) | 1658 (50.1%) |  |
| Neonatal weight (gram) | 3310.0 [2990.0;3630.0] | 3240.0 [2860.0;3570.0] | 3300.0 [2970.0;3630.0] | **<0.001** |
| Macrosomia (>4000 g) | 247 ( 8.6%) | 30 ( 7.0%) | 277 ( 8.4%) | 0.33 |
| IUGR | 121 ( 4.2%) | 30 ( 7.0%) | 151 ( 4.6%) | **0.01** |
| Neonatal length (cm) | 50.5 [49.0;52.0] | 50.0 [49.0;52.0] | 50.5 [49.0;52.0] | **<0.001** |
| Head circumference (cm) | 34.0 [33.0;35.0] | 34.0 [33.0;35.0] | 34.0 [33.0;35.0] | 0.12 |
| Placental weight (gram) | 466.0 [404.0;539.0] | 456.0 [398.0;534.5] | 466.0 [403.0;538.0] | 0.30 |
| Fetal placental ratio (FPR) | 7.1 [ 6.3; 7.8] | 7.0 [ 6.3; 7.7] | 7.1 [ 6.3; 7.8] | 0.28 |
| Cord length | 36.0 [28.0;43.0] | 34.0 [27.0;42.0] | 35.0 [28.0;43.0] | 0.14 |
| Coiling | 4.0 [ 3.0; 5.0] | 4.0 [ 3.0; 5.0] | 4.0 [ 3.0; 5.0] | 0.99 |
| **Maternal characteristics and complications** | | | | |
| Race /ethnicity |  |  |  | 0.60 |
| - Asian | 127 ( 4.4%) | 22 ( 5.2%) | 149 ( 4.5%) |  |
| - Black | 947 (32.9%) | 150 (35.1%) | 1097 (33.2%) |  |
| - Hispanic | 241 ( 8.4%) | 38 ( 8.9%) | 279 ( 8.4%) |  |
| - Others/unknown/declined | 223 ( 7.7%) | 26 ( 6.1%) | 249 ( 7.5%) |  |
| - White | 1342 (46.6%) | 191 (44.7%) | 1533 (46.4%) |  |
| Marital status |  |  |  | **<0.001** |
| - Divorced | 13 ( 0.5%) | 2 ( 0.5%) | 15 ( 0.5%) |  |
| - Life partner | 216 ( 7.5%) | 59 (13.8%) | 275 ( 8.3%) |  |
| - Married | 1706 (59.2%) | 239 (56.0%) | 1945 (58.8%) |  |
| - Others/Unknown/Declined | 18 ( 0.6%) | 0 ( 0.0%) | 18 ( 0.5%) |  |
| - Single | 927 (32.2%) | 127 (29.7%) | 1054 (31.9%) |  |
| Maternal age (year) | 32.0 [27.0;36.0] | 32.0 [27.0;36.0] | 32.0 [27.0;36.0] | 0.72 |
| Gestational age (week) | 40.0 [39.0;40.0] | 39.0 [38.0;40.0] | 39.0 [39.0;40.0] |  |
| BMI at delivery | 30.7 [27.6;35.5] | 31.1 [27.4;35.5] | 30.8 [27.6;35.5] | 0.91 |
| Maternal obesity (BMI >30) | 1077 (55.7%) | 164 (56.6%) | 1241 (55.8%) | 0.84 |
| Maternal obnesity class |  |  |  | 0.95 |
| - No obesity (BMI<30) | 856 (44.3%) | 126 (43.4%) | 982 (44.2%) |  |
| - Obesity class I (BMI 30-34) | 550 (28.5%) | 86 (29.7%) | 636 (28.6%) |  |
| - Obesity class II (BMI 35 -39) | 305 (15.8%) | 47 (16.2%) | 352 (15.8%) |  |
| - Obesity class III (BMI 40 or over) | 222 (11.5%) | 31 (10.7%) | 253 (11.4%) |  |
| Delivery |  |  |  | 0.70 |
| - Cesarean | 996 (34.6%) | 143 (33.5%) | 1139 (34.4%) |  |
| - Vaginal | 1884 (65.4%) | 284 (66.5%) | 2168 (65.6%) |  |
| GBS status | 456 (31.6%) | 62 (30.4%) | 518 (31.5%) | 0.78 |
| SARS-CoV2 status | 165 ( 5.7%) | 22 ( 5.2%) | 187 ( 5.7%) | 0.72 |
| **PRE/PIH** | **450 (15.6%)** | **43 (10.1%)** | **493 (14.9%)** | **<0.001** |
| IUFD | 6 ( 0.2%) | 0 ( 0.0%) | 6 ( 0.2%) | 0.74 |
| IUGR | 121 ( 4.2%) | 30 ( 7.0%) | 151 ( 4.6%) | **0.01** |
| Category 2 fetal tracing | 604 (21.0%) | 78 (18.3%) | 682 (20.6%) | 0.22 |
| Oligohydramnios | 68 ( 2.4%) | 6 ( 1.4%) | 74 ( 2.2%) | 0.28 |
| **Placental pathology** | | | | |
| **Maternal vascular malperfusion (MVM)** | |  |  |  |
| Decidual vasculopathy |  |  |  | 1.00 |
| - Classic type | 743 (25.8%) | 109 (25.5%) | 852 (25.8%) | 0.95 |
| - Mixed type | 134 ( 4.7%) | 20 ( 4.7%) | 154 ( 4.7%) | 1.00 |
| - Mural type | 229 ( 8.0%) | 35 ( 8.2%) | 264 ( 8.0%) | 0.94 |
| Placental infarcts | 191 ( 6.6%) | 21 ( 4.9%) | 212 ( 6.4%) | 0.21 |
| Maternal thrombosis | 577 (20.0%) | 88 (20.6%) | 665 (20.1%) | 0.83 |
| Placental abruption | 45 ( 1.6%) | 2 ( 0.5%) | 47 ( 1.4%) | 0.12 |
| **Fetal vascular malperfusion (FVM)** | 353 (12.3%) | 48 (11.2%) | 401 (12.1%) | 0.60 |
| **Inflammatory/infectious** |  |  |  |  |
| Acute chorioamnionitis (MIR) | 983 (34.1%) | 141 (33.0%) | 1124 (34.0%) | 0.69 |
| Chronic deciduitis (>50/HPF)(MIR) | 735 (25.5%) | 101 (23.7%) | 836 (25.3%) | 0.44 |
| Chronic villitis (MIR) | 581 (20.2%) | 88 (20.6%) | 669 (20.2%) | 0.89 |
| Acute funisitis /fetal vasculitis (FIR) | 389 (13.5%) | 50 (11.7%) | 439 (13.3%) | 0.35 |
| **Other placental pathology** |  |  |  |  |
| Meconium stain | 919 (31.9%) | 121 (28.3%) | 1040 (31.5%) | 0.15 |
| Subchorionic hematoma (> 1.0 cm) | 232 ( 8.1%) | 37 ( 8.7%) | 269 ( 8.1%) | 0.74 |
| MPFD/MFI | 88 ( 3.1%) | 15 ( 3.5%) | 103 ( 3.1%) | 0.72 |
| **Lab and other tests** |  |  |  |  |
| WBC count (x1000) | 9.7 [ 8.2;11.8] | 9.7 [ 8.1;11.7] | 9.7 [ 8.2;11.8] | 0.37 |
| Neutrophil differential (%) | 72.5 [67.8;77.3] | 72.5 [67.5;78.0] | 72.5 [67.7;77.4] | 0.72 |
| Lymphocyte differential (%) | 17.9 [14.2;22.2] | 17.7 [13.7;22.1] | 17.9 [14.2;22.2] | 0.50 |
| Body temperature (^0^C) | 36.7 [36.5;37.0] | 36.7 [36.5;36.9] | 36.7 [36.5;37.0] | 0.56 |
| Blood pressure (Systolic) | 126.0 [118.0;135.0] | 123.0 [114.0;134.0] | 126.0 [117.0;135.0] |  |
| Blood pressure (Diastolic) | 78.0 [71.0;84.0] | 77.0 [70.0;83.0] | 77.0 [70.0;84.0] |  |
| Abbreviation: PRE - preeclampsia, PIH - pregnancy induced hypertension; GBS - group B streptococcus; GDM2 - | | | | |
| gestational diabetes mellitus; IUGR - intrauterine growth restriction; IUFD - intrauterine fetal demise; | | | | |
| MPFD/ MFI - massive perivillous fibrinoid deposit /maternal floor infarction; BMI - body mass index. | | | |  |
| Values expressed were mean and percentage with 95% confidence intervals. P<0.05 is considered significant. | | | | |

| **Supplementary table 3: Clinical and placental pathology features of combined preeclampsia and diabetes** | | | | | | |
| --- | --- | --- | --- | --- | --- | --- |
|  | **None** | **GDM** | **PRE/PIH/GDM2** | **PRE/PIH** | **Total** | **p value** |
|  | **(N=2713)** | **(N=397)** | **(N=55)** | **(N=559)** | **(N=3724)** |  |
| **Neonatal birth data** | | | | | | |
| Neonatal sex |  |  |  |  |  | 0.77 |
| - Female | 1327 (48.9%) | 192 (48.4%) | 29 (52.7%) | 284 (50.8%) | 1832 (49.2%) |  |
| - Male | 1375 (50.7%) | 205 (51.6%) | 26 (47.3%) | 272 (48.7%) | 1878 (50.4%) |  |
| Neonatal weight (gram) | 3250.0 [2870.0;3600.0] | 3350.0 [3010.0;3670.0] | 3300.0 [2887.5;3595.0] | 3000.0 [2565.0;3390.0] | 3230.0 [2840.0;3580.0] | **<0.001** |
| Macrosomia (>=4000 g) | 194 ( 7.2%) | 44 (11.1%) | 9 (16.4%) | 31 ( 5.6%) | 278 ( 7.5%) | **<0.001** |
| Neonatal length (cm) | 50.0 [48.5;52.0] | 50.0 [49.0;52.0] | 50.0 [48.2;52.0] | 49.5 [47.0;51.0] | 50.0 [48.0;52.0] |  |
| Neonatal head circumference (cm) | 34.0 [33.0;35.0] | 34.0 [33.0;35.0] | 34.0 [33.0;35.0] | 33.5 [32.0;34.5] | 34.0 [33.0;35.0] |  |
| Placental weight (gram) | 454.0 [390.0;528.0] | 478.0 [415.0;556.0] | 486.0 [399.0;589.5] | 432.0 [365.5;511.5] | 454.0 [389.0;530.0] | **<0.001** |
| Fetal placental ratio (FPR) | 7.0 [ 6.2; 7.8] | 6.9 [ 6.2; 7.8] | 6.3 [ 6.0; 7.2] | 6.7 [ 5.9; 7.6] | 7.0 [ 6.2; 7.8] | **<0.001** |
| Umbilical cord length (cm) | 35.0 [27.0;42.0] | 34.0 [26.0;42.0] | 41.0 [29.0;48.0] | 34.0 [26.0;42.0] | 35.0 [27.0;42.0] | **0.03** |
| Umbilical cord coiling per 10 cm | 4.0 [ 3.0; 5.0] | 4.0 [ 3.0; 5.0] | 3.0 [ 3.0; 5.0] | 4.0 [ 3.0; 5.0] | 4.0 [ 3.0; 5.0] | 0.71 |
| **Maternal characteristics and complications** | | | | | | |
| Race/ethnicity |  |  |  |  |  | **<0.001** |
| - Asian | 101 ( 3.7%) | 36 ( 9.1%) | 6 (10.9%) | 12 ( 2.1%) | 155 ( 4.2%) |  |
| - Black | 880 (32.4%) | 104 (26.2%) | 24 (43.6%) | 283 (50.6%) | 1291 (34.7%) |  |
| - Hispanic | 214 ( 7.9%) | 47 (11.8%) | 6 (10.9%) | 48 ( 8.6%) | 315 ( 8.5%) |  |
| - Others/Unknown/Declined | 226 ( 8.3%) | 36 ( 9.1%) | 5 ( 9.1%) | 34 ( 6.1%) | 301 ( 8.1%) |  |
| - White | 1292 (47.6%) | 174 (43.8%) | 14 (25.5%) | 182 (32.6%) | 1662 (44.6%) |  |
| Marital status |  |  |  |  |  | **<0.001** |
| - Divorced | 9 ( 0.3%) | 5 ( 1.3%) | 0 ( 0.0%) | 2 ( 0.4%) | 16 ( 0.4%) |  |
| - Life partner | 213 ( 7.9%) | 32 ( 8.1%) | 4 ( 7.3%) | 63 (11.3%) | 312 ( 8.4%) |  |
| - Married | 1607 (59.2%) | 247 (62.2%) | 28 (50.9%) | 267 (47.8%) | 2149 (57.7%) |  |
| - Others/Unknown/Declined | 7 ( 0.3%) | 0 ( 0.0%) | 0 ( 0.0%) | 1 ( 0.2%) | 8 ( 0.2%) |  |
| - Single | 867 (32.0%) | 113 (28.5%) | 23 (41.8%) | 224 (40.1%) | 1227 (32.9%) |  |
| Delivery mode |  |  |  |  |  | **<0.001** |
| - Cesarean | 897 (33.1%) | 148 (37.3%) | 28 (50.9%) | 240 (42.9%) | 1313 (35.3%) |  |
| - Vaginal | 1816 (66.9%) | 249 (62.7%) | 27 (49.1%) | 319 (57.1%) | 2411 (64.7%) |  |
| Maternal age (year) | 31.0 [27.0;35.0] | 34.0 [29.0;37.0] | 35.0 [31.0;38.0] | 32.0 [28.0;36.0] | 32.0 [27.0;36.0] | **<0.001** |
| Gestational age (week) | 40.0 [38.0;40.0] | 39.0 [38.0;40.0] | 38.0 [37.0;39.0] | 38.0 [37.0;39.0] | 39.0 [38.0;40.0] |  |
| Preterm delivery (<37 week) | 282 (10.4%) | 13 ( 3.3%) | 12 (21.8%) | 109 (19.5%) | 416 (11.2%) | **<0.001** |
| Body mass index at delivery (BMI) | 29.9 [26.7;34.4] | 32.4 [28.4;37.1] | 36.5 [31.4;42.2] | 34.0 [29.7;38.2] | 30.8 [27.4;35.6] | **<0.001** |
| Maternal obesity (BMI>30) | 892 (49.9%) | 191 (65.2%) | 32 (80.0%) | 296 (73.3%) | 1411 (55.9%) | **<0.001** |
| Obesity classes |  |  |  |  |  | **<0.001** |
| - No obesity (BMI<30) | 896 (50.1%) | 102 (34.8%) | 8 (20.0%) | 108 (26.7%) | 1114 (44.1%) |  |
| - Obesity class I (BMI 30 - 34) | 494 (27.6%) | 98 (33.4%) | 10 (25.0%) | 118 (29.2%) | 720 (28.5%) |  |
| - Obesity class II (BMI 35 - 39) | 232 (13.0%) | 54 (18.4%) | 8 (20.0%) | 100 (24.8%) | 394 (15.6%) |  |
| - Obesity class III (BMI 40 or over) | 166 ( 9.3%) | 39 (13.3%) | 14 (35.0%) | 78 (19.3%) | 297 (11.8%) |  |
| GBS status | 384 (30.7%) | 68 (32.7%) | 8 (28.6%) | 92 (36.2%) | 552 (31.7%) | 0.36 |
| SARS-CoV2 status | 180 ( 6.6%) | 9 ( 2.3%) | 1 ( 1.8%) | 9 ( 1.6%) | 199 ( 5.3%) | **<0.001** |
| PRE/PIH | 0 ( 0.0%) | 0 ( 0.0%) | 55 (100.0%) | 559 (100.0%) | 614 (16.5%) |  |
| GDM2 | 0 ( 0.0%) | 397 (100.0%) | 55 (100.0%) | 0 ( 0.0%) | 452 (12.1%) |  |
| IUGR | 138 ( 5.1%) | 15 ( 3.8%) | 0 ( 0.0%) | 27 ( 4.8%) | 180 ( 4.8%) | 0.25 |
| IUFD | 41 ( 1.5%) | 0 ( 0.0%) | 0 ( 0.0%) | 7 ( 1.3%) | 48 ( 1.3%) | 0.07 |
| Category 2 fetal tracing | 653 (24.1%) | 24 ( 6.0%) | 4 ( 7.3%) | 32 ( 5.7%) | 713 (19.1%) | **<0.001** |
| Oligohydramnios | 83 ( 3.1%) | 4 ( 1.0%) | 0 ( 0.0%) | 0 ( 0.0%) | 87 ( 2.3%) | **<0.001** |
| **Placental pathology** | | | | | | |
| **Maternal vascular malperfusion (MVM)** | |  |  |  |  |  |
| Decidual vasculopathy |  |  |  |  |  |  |
| - Classic type | 703 (25.9%) | 91 (22.9%) | 14 (25.5%) | 167 (29.9%) | 975 (26.2%) | 0.10 |
| - Mixed type | 116 ( 4.3%) | 17 ( 4.3%) | 10 (18.2%) | 70 (12.5%) | 213 ( 5.7%) | **<0.001** |
| - Mural arterial hypertrophy | 163 ( 6.0%) | 48 (12.1%) | 6 (10.9%) | 71 (12.7%) | 288 ( 7.7%) | **<0.001** |
| Placental infarcts | 153 ( 5.6%) | 21 ( 5.3%) | 4 ( 7.3%) | 94 (16.8%) | 272 ( 7.3%) | **<0.001** |
| Maternal thrombosis | 535 (19.7%) | 82 (20.7%) | 11 (20.0%) | 135 (24.2%) | 763 (20.5%) | 0.13 |
| Placental abruption | 42 ( 1.5%) | 1 ( 0.3%) | 3 ( 5.5%) | 21 ( 3.8%) | 67 ( 1.8%) | **<0.001** |
| **Fetal vascular malperfusion (FVM)** | |  |  |  |  |  |
| - Avascular villi | 332 (12.2%) | 50 (12.6%) | 4 ( 7.3%) | 52 ( 9.3%) | 438 (11.8%) | 0.16 |
| **Inflammatory/Infectious** |  |  |  |  |  |  |
| Acute chorioamnionitis (MIR) | 1042 (38.4%) | 98 (24.7%) | 15 (27.3%) | 110 (19.7%) | 1265 (34.0%) | **<0.001** |
| Chronic deciduitis (>50/PHF)(MIR) | 696 (25.7%) | 106 (26.7%) | 13 (23.6%) | 101 (18.1%) | 916 (24.6%) | **<0.001** |
| Chronic villitis (MIR) | 544 (20.1%) | 96 (24.2%) | 12 (21.8%) | 75 (13.4%) | 727 (19.5%) | **<0.001** |
| Acute funisitis/fetal vasculitis (FIR) | 422 (15.6%) | 39 ( 9.8%) | 6 (10.9%) | 34 ( 6.1%) | 501 (13.5%) | **<0.001** |
| **Other placental pathology** |  |  |  |  |  |  |
| Meconium stain fetal membranes | 936 (34.5%) | 65 (16.4%) | 2 ( 3.6%) | 61 (10.9%) | 1064 (28.6%) | **<0.001** |
| Subchorionic hematoma (>1.0 cm) | 242 ( 8.9%) | 30 ( 7.6%) | 3 ( 5.5%) | 25 ( 4.5%) | 300 ( 8.1%) | **0.01** |
| **Lab and other tests** |  |  |  |  |  |  |
| WBC count (x1000) | 10.0 [ 8.3;12.3] | 9.4 [ 7.8;11.1] | 8.8 [ 7.1;11.0] | 9.4 [ 7.9;11.1] | 9.8 [ 8.2;11.9] |  |
| Neutrophil differential (%) | 73.0 [67.9;78.0] | 71.6 [67.5;75.9] | 70.1 [65.4;75.9] | 70.9 [66.0;75.8] | 72.5 [67.6;77.4] |  |
| Lymphocyte differential (%) | 17.5 [13.7;21.7] | 19.2 [15.3;23.2] | 20.0 [14.8;24.5] | 19.1 [15.2;24.1] | 18.0 [14.2;22.3] |  |
| Body temperature (^0^C) | 36.7 [36.5;37.0] | 36.7 [36.5;37.0] | 36.7 [36.6;37.0] | 36.7 [36.5;37.0] | 36.7 [36.5;37.0] | 0.34 |
| Blood pressure (Systolic) | 123.0 [116.0;131.0] | 125.0 [118.5;132.0] | 143.0 [137.0;148.0] | 145.0 [138.0;152.0] | 126.0 [118.0;136.0] | **<0.001** |
| Blood pressure (Diastolic) | 75.0 [69.0;81.0] | 77.0 [71.0;82.0] | 90.0 [84.0;95.0] | 89.0 [83.5;96.0] | 78.0 [70.0;84.0] | **<0.001** |
| Abbreviation: PRE - preeclampsia, PIH - pregnancy induced hypertension; GBS - group B streptococcus; GDM2 - gestational diabetes mellitus; | | | | | | |
| IUGR - intrauterine growth restriction; IUFD - intrauterine fetal demise; MPFD/ MFI - massive perivillous fibrinoid deposit /maternal floor infarction; | | | | | | |
| BMI - body mass index. Values expressed were mean and percentage with 95% confidence intervals. P<0.05 is considered significant. | | | | | |  |

S Figure 1 Logistic regression model of GDM2 and total pregnancy

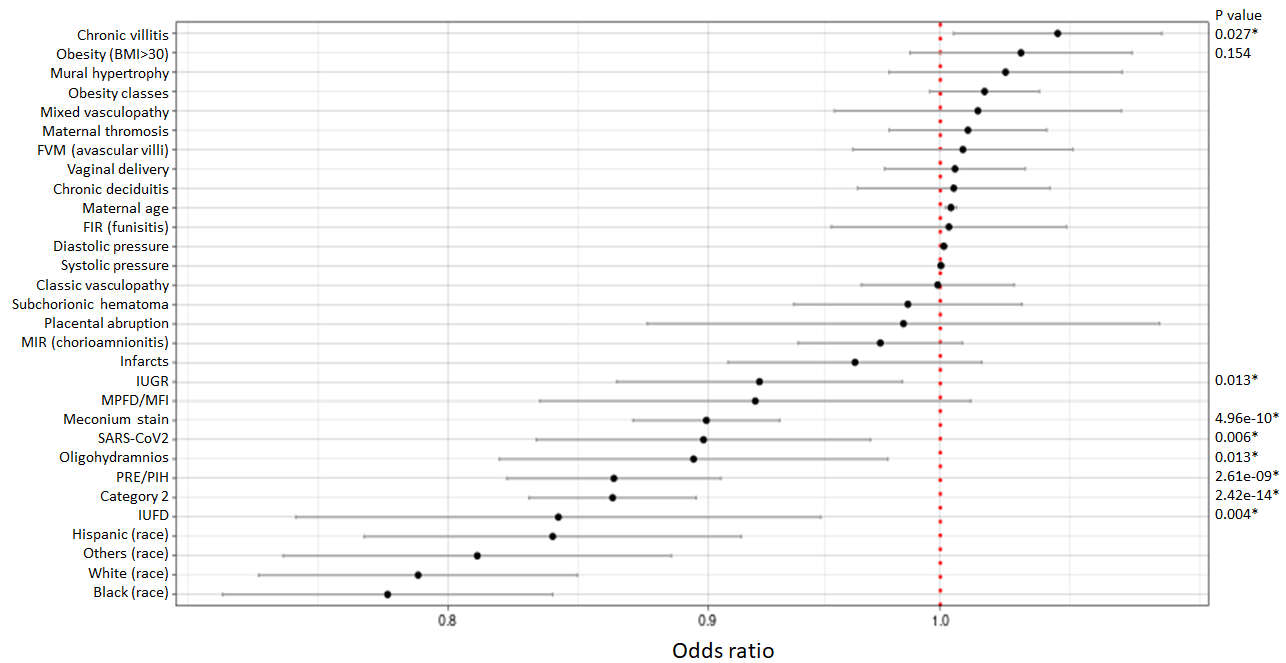

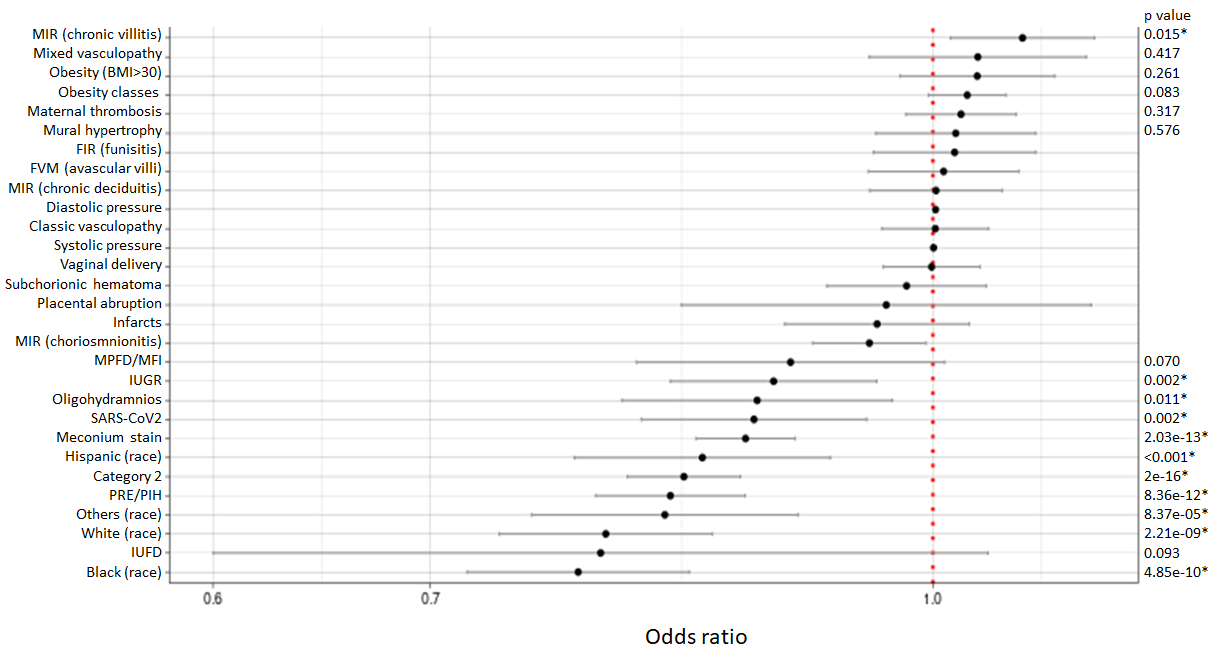

S Figure 2 Logistic regression model of GDM2 and term pregnancy
